# Low Back Pain’s Hidden Partners: Stigma, Anxiety, and Functional Decline in Adults with Cerebral Palsy

**DOI:** 10.1101/2024.12.07.24318657

**Authors:** Julie A. Stutzbach, Cristina A. Sarmiento, Tanya S. Kenkre, Joyce L. Oleszek, Stephen R Wisniewski, Mary E. Gannotti, Cerebral Palsy Research Network

## Abstract

**AIM:** Determine biopsychosocial factors associated with pain interference and pain intensity in adults with cerebral palsy (CP) and chronic low back pain (LBP).

**METHOD:** Cross-sectional secondary data analysis of a community survey examining function and chronic pain in adults with CP and LBP. We examined bivariate relationships and built two regression models with pain interference with general activities and pain intensity as dependent variables and biopsychosocial factors as explanatory factors.

**RESULTS:** We included 295 participants with CP and LBP in our analyses. Average age was 43.2 ± 13.9 years, and 81% were ambulatory (GMFCS I-III). Pain was present for 17.9 ± 13.4 years. Ordinary least squares regression models indicated greater pain interference with: change in best motor function since childhood (p=0.002), stigma (p=0.01), and anxiety (p=0.01; N=238; adjusted R^2^=0.17); and greater pain intensity with: lower income (p=0.01), Hispanic/Latino ethnicity (p=0.04), anxiety (p=0.01), and less satisfaction with social roles, (p<0.001; N=290, R^2^=0.18; Adjusted R^2^=0.16).

**INTERPRETATION:** These findings emphasize the importance of assessing and interpreting chronic pain in the context of biopsychosocial factors, particularly anxiety, stigma, race, ethnicity, income, satisfaction with social roles, and changes in motor function.

## Introduction

Sequelae of cerebral palsy (CP) range from minor disability to complete dependence on caregivers for activities of daily living.(1) As a highly heterogeneous condition, CP can cause a variety of impairments including weakness, movement disorders, communication difficulties, gait deviations, reduced motor control, intellectual disability, and – importantly – pain.(2) While CP is not a progressive condition, many individuals with CP undergo a precipitous decline in function and health-related quality of life during their transition from childhood to adulthood,(3) in part because of chronic pain.(4) Up to seventy percent of adults with CP experience chronic pain,(5) which is one of the main reasons for declines in ambulation and other functional activities.(6) More specifically, low back pain (LBP) is the most common physical location for pain among adults with CP and chronic pain.(7)

Pain’s impact on functioning (i.e., pain interference) is a critical construct of chronic pain, as it captures pain-related disability.(8) Studies suggest high variability in pain interference among individuals with CP, with some describing minimal disability(6,9,10) while others finding moderate to high pain interference.(11,12) Investigators have postulated that lower pain interference levels may come from coping strategies related to living with a chronic disability or lower baseline functional status stemming from motor and cognitive symptoms.(8) Despite these hypotheses, the reasons why some adults with CP are susceptible to greater pain interference remain unknown.(8)

Social determinants of health likely play a role in pain disparities for people with CP. Among people without CP who have LBP, racial and ethnic differences in perceptions of pain intensity, interference, and access to health care are well documented.(13) The Institute of Medicine identifies several risk factors for pain and poor pain treatment including, English as a second language, low income and education, and woman gender.(14) However, the relationship between these biopsychosocial factors and LBP in adults with CP has not been well described. This study will use the biopsychosocial model, which provides a comprehensive picture of the biological, social/environmental, and psychological factors that may all play a role in the development and persistence of chronic pain.(15)

A plethora of research supports an integrated care model for children with CP that includes highly specialized, interprofessional care.(16) However, this care model tends to dissolve after the transition to adult-based care,(17) making adults with CP an underserved and under resourced population. Adults with CP are less likely than the general population to receive treatment for their pain,(18) such as physical therapy,(19,20) even though they might have a greater need for interprofessional pain care than the general population. There are many reasons for underutilization of services, and provider knowledge about aging and neurodevelopmental disabilities is a contributing factor.(21) An improved understanding of adults with CP and their LBP can lead to more rapid translation of strategies to mitigate this disabling issue. A better understanding of pain interference and intensity, and associated factors, could improve pain care across the spectrum of associated disability.

The aim of this study was to identify personal and social factors that influence pain severity and pain interference in adults with CP and chronic LBP. We had three hypotheses: 1) greater stigma would be associated with increased pain interference (as stigma is a barrier to seeking care and adds to the burden of chronic pain), 2) those who had experienced pain for a longer duration would have less pain interference (due to development of resilience to pain and coping strategies), and 3) individuals who are non-ambulatory will have less pain interference than ambulatory adults with CP due to lower baseline functional status. We also conducted an exploratory analysis of additional social determinants of health such as income, ethnicity, and race; psychological factors such as anxiety and depression; and physical factors such as decline in gross motor function that could be associated with pain interference and pain intensity.

## Methods

This was a cross-sectional, secondary data analysis of an ongoing Cerebral Palsy Research Network (CPRN) Community Registry study of adults with CP(22) which collects self-reported measures of function, pain, and potential related factors. The parent study was developed in conjunction with stakeholders(23) including people with CP, caregivers, clinicians, and researchers in a process outlined elsewhere.(24) Participants are eligible to participate in the Community Registry study if they self-identify as having CP, are at least 18 years old, are able to understand English, and can provide informed consent. We pulled data from a subsample of the participants who identified the presence of chronic pain (defined as pain that is persistent for >3 months)(25) and indicated that they had pain in the lower back. All participants who met these criteria were included in the analysis.

The Cerebral Palsy Research Network (CPRN) Community Registry data collection is approved by the University of Pittsburgh Institutional Review Board Institutional Review Board (IRB) Participants were recruited into the registry from social media, webinars, and at conferences, in a process outlined elsewhere.(26) All Community Registry participants provide informed consent.

### Statistical analysis

#### Outcome measures and explanatory factors

All measures in the parent study are based on self-report. Our primary outcome measures were two components of the brief pain inventory(27), both on a 0-10 numerical rating scale: pain intensity (average of preceding 24 hours) and pain interference with general activities (over the preceding 24 hours).

Potential explanatory factors included ambulation status as indicated by Gross Motor Function Classification System level(28) (GMFCS; dichotomized into ambulatory (levels I-III) vs. non-ambulatory (levels IV-V)); gender (female vs. male); race (White vs. African American vs. Other race); ethnicity (Hispanic/Latino vs. Non-Hispanic/Latino); education level (up to Associate degree vs. Bachelor degree/graduate/professional degree); current paid employment (yes vs. no); income above or below $41,998;(29) current age; years since onset of pain; and whether or not there has been change in best motor function since childhood. We also included various validated measures as collected by the parent study to capture psychosocial outcomes, including: Quality of Life in Neurological Disorders (Neuro-QoL) Stigma;(30) Patient-Reported Outcomes Measurement Information System (PROMIS) Anxiety;(31) PROMIS Depression; (32) PROMIS Satisfaction with Roles and Activities(33). PROMIS and NeuroQoL scores are transformed into a final T-score (mean 50, ± 10) based on normative data, with higher scores indicating more of the domain measured (e.g., higher score = more stigma, more depression).

#### Associations with outcomes and multivariate regression models

First, we evaluated bivariate relationships between variables of interest and primary outcomes (pain interference and pain intensity) and pain interference/intensity with each other with the T-test or Wilcoxon test (dichotomous measures); ANOVA (categorical measures); and Pearson correlation test (continuous measures).

We used ordinary least squares regression to build multivariate regression models for pain interference and pain intensity. We followed an iterative approach and evaluated multiple combinations of the biopsychosocial factors that demonstrated alpha ≤0.20 in bivariate association with pain outcomes. We eliminated variables with alpha > 0.10 in a multivariate model if the parameter estimates of the remaining variables did not change by more than 20% after eliminating the variable.(34) After several iterations, we were left with a core set of explanatory variables. We then added back into the model each variable that had been eliminated (retained if significant), and confirmed that the alpha values of the core set of explanatory variables remained significant at the 0.05 level and that their parameter estimates remained stable. We additionally examined variance inflation factors to determine that there was not multicollinearity among the variables included in the final model. Thus, the variables retained in the final models are those that were consistently significantly associated with pain interference and of pain intensity and that demonstrated the most stability in various combinations of covariates. (34) We did not make corrections for multiple tests due to the exploratory nature of this study.

Statistical significance for all analyses was set at alpha <0.05. SAS v9.4 (©2016, Cary, NC USA) was used for analyses.

## Results

### Timeline

Data collection began in April 2019 and is ongoing. For this analysis, data from the registry were pulled in November of 2023. 996 adults with CP signed up for the registry, and 695 respondents consented to respond to surveys about functional change and chronic pain. Of the 572 who identified chronic pain and completed the chronic pain surveys, 457 identified presence of low back pain. We excluded 162 respondents who were missing responses for PROMIS Anxiety measures and/or the measure of years since onset of pain as these variables were integral to our research question. 295 respondents were included in this analysis. We compared the survey responses on the measures we evaluated for associations with and as multivariate predictors of pain outcomes between the two groups (295 included in analyses vs. 162 excluded from analyses) and found no statistically significant differences in the distribution of responses between the two groups (alpha for significance < 0.05).

### Subject characteristics

Table 1 depicts demographics and clinical characteristics including, GMFCS level, education, employment, income, years since pain onset, age, and change in best motor function since childhood. Participants were majority female and ambulatory, and ranged in age from 18.3 to 77.9 years. Their pain had been present, on average, for almost 18 years. Average pain intensity and pain interference were both moderate (5.2/10 and 4.3/10, respectively), though both ranged from 0-10.

**Table 1.**
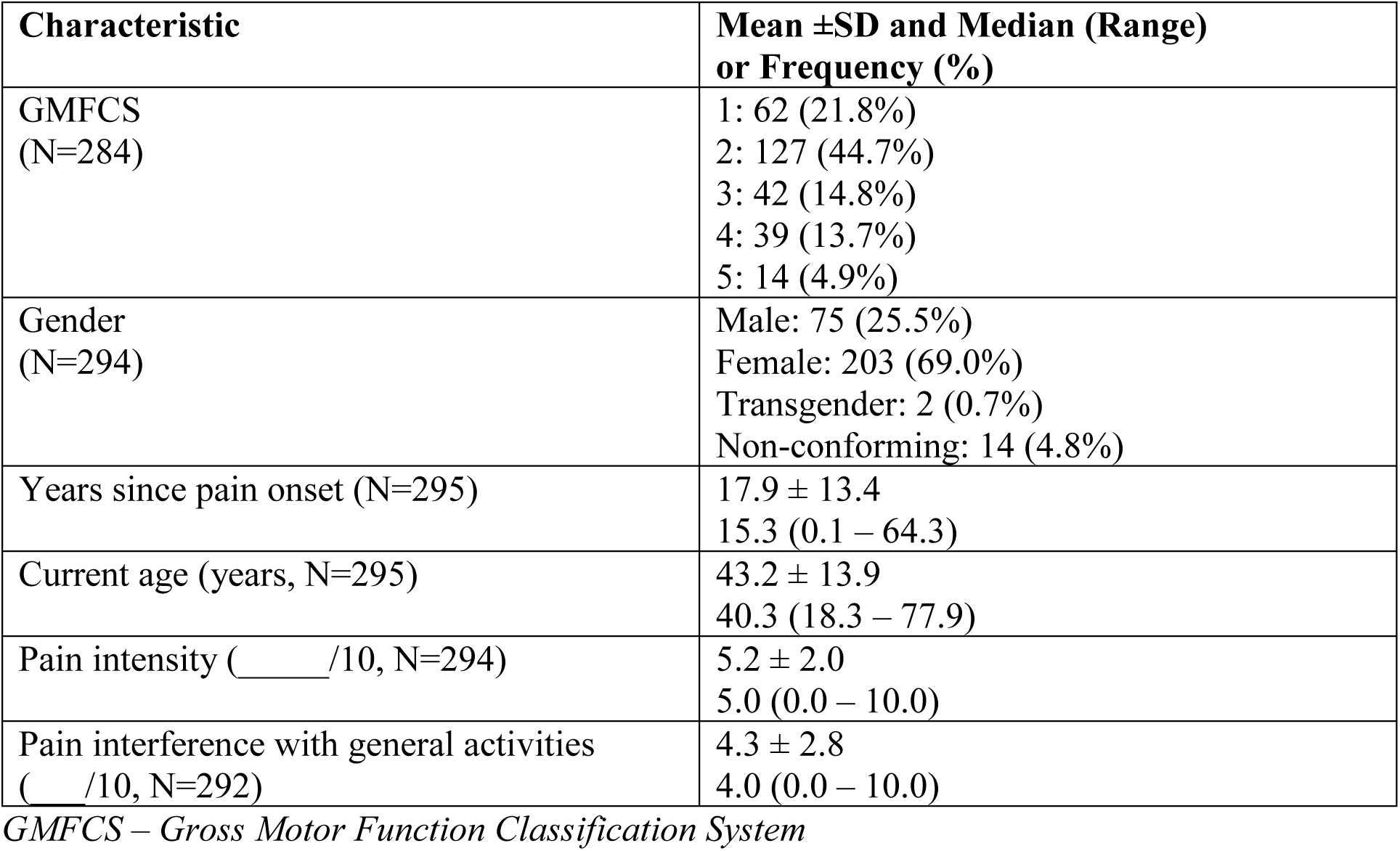
Demographics and clinical characteristics of population.

| Characteristic | Mean ±SD and Median (Range) or Frequency (%) |
| --- | --- |
| GMFCS<br>(N=284) | 1: 62 (21.8%)<br>2: 127 (44.7%)<br>3: 42 (14.8%)<br>4: 39 (13.7%)<br>5: 14 (4.9%) |
| Gender<br>(N=294) | Male: 75 (25.5%)<br>Female: 203 (69.0%)<br>Transgender: 2 (0.7%)<br>Non-conforming: 14 (4.8%) |
| Years since pain onset (N=295) | 17.9 ± 13.4<br>15.3 (0.1 – 64.3) |
| Current age (years, N=295) | 43.2 ± 13.9<br>40.3 (18.3 – 77.9) |
| Pain intensity (____/10, N=294) | 5.2 ± 2.0<br>5.0 (0.0 – 10.0) |
| Pain interference with general activities<br>(____/10, N=292) | 4.3 ± 2.8<br>4.0 (0.0 – 10.0) |

*GMFCS – Gross Motor Function Classification System*

### Statistical analyses

Table 2 shows the bivariate relationships between biopsychosocial variables and pain interference/pain intensity. Pain intensity and interference were significantly correlated (ρ = 0.60, P < 0.0001). Figure 1A and 1B show variables tested nested within the biopsychosocial model of pain, if they were associated with primary outcomes, and if they were included in the final models. The following variables were significantly associated with higher pain interference: no current paid employment; greater levels of stigma, anxiety, and depression; lower levels of satisfaction with social roles and activities; and change in best motor function since childhood.

**Figure 1:**
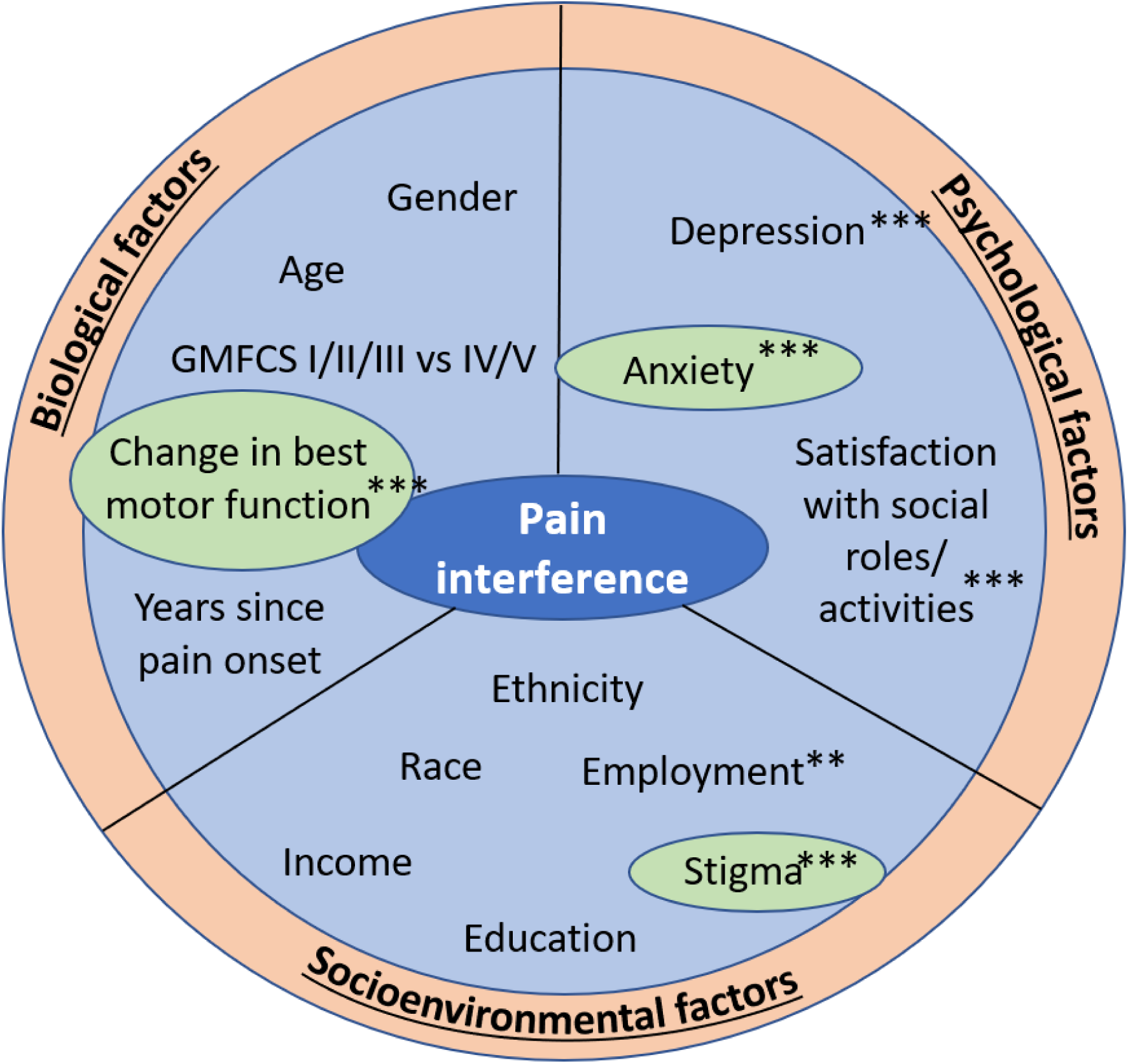
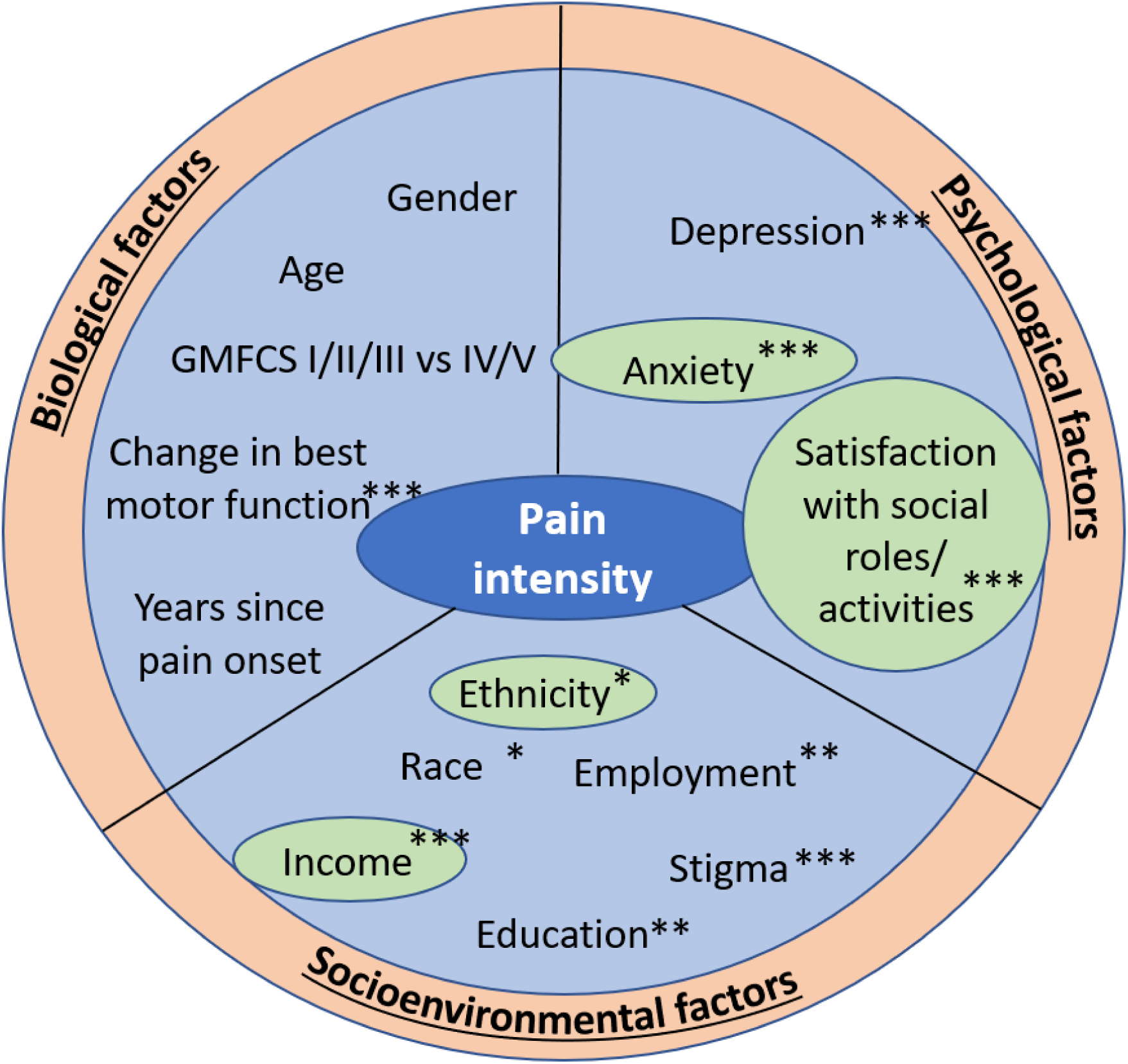
Factors associated with pain interference (Figure 1A) and pain intensity (Figure 1B) in adults with CP organized by the biopsychosocial model of pain *significant bivariate association and candidate for final model (*P<.05, **P<.01, ***P<.001) Green/Circled factors were included in final model

**Table 2.** Bivariate associations between biopsychosocial measures and pain outcomes. P-values listed are those for applicable tests of association (T-test, Wilcoxon test, ANOVA, or Pearson correlation test); for Pearson correlation tests, the rho value (ρ) is also listed.

| Variable | Mean ± SD (range) or counts (%) | Pain Interference | Pain Intensity |
| --- | --- | --- | --- |
| Current age (N=295) | 43.2 ± 13.9 (18.3 – 77.9) | $\rho$ = 0.05<br>p-value = 0.40 | $\rho$ = -0.03<br>p-value = 0.62 |
| Ambulatory status (N=284) | GMFCS I-III (ambulatory): 231 (81.3%)<br>GMFCS IV-V (non-ambulatory): 53 (18.7%) | 4.2 ± 2.8 (0.0 -10.0)<br>4.3 ± 2.7 (0.0 -10.0)<br>p-value = 0.84 | 5.1 ± 2.0 (0.0 – 10.0)<br>5.6 ± 2.0 (1.0 – 10.0)<br>p-value = 0.09 |
| Gender (N=278) | Male: 75 (27.0%)<br>Female: 203 (73.0%) | 3.9 ± 2.8 (0.0 -10.0)<br>4.3 ± 2.7 (0.0 -10.0)<br>p-value = 0.27 | 4.8 ± 2.1 (0.0 – 9.0)<br>5.3 ± 2.0 (0.0 – 10.0)<br>p-value = 0.08 |
| Race (N=287) | White: 266 (92.7%)<br>African American: 10 (3.5%)<br>Other: 11 (3.8%) | 4.2 ± 2.7 (0.0 -10.0)<br>4.9 ± 3.8 (0.0 -10.0)<br>4.6 ± 3.5 (0.0 -10.0)<br>p-value = 0.67 | 5.1 ± 2.0 (0.0 – 10.0)<br>6.7 ± 1.6 (5.0 – 10.0)<br>4.7 ± 2.1 (1.0 – 8.0)<br>p-value = 0.04* |
| Ethnicity (N=295) | Hisp/Lat: 13 (4.4%)<br>Not Hisp/Lat: 282 (95.6%) | 4.3 ± 2.6 (0.0 -8.0)<br>4.3 ± 2.8 (0.0 -10.0)<br>p-value = 0.89 | 6.2 ± 2.2 (1.0 – 8.0)<br>5.2 ± 2.0 (0.0 – 10.0)<br>p-value = 0.03* |
| Education (N=294) | ≤ Associate degree: 111 (37.8%)<br>Bachelor or Grad/Prof: 183 (62.2%) | 4.7 ± 2.9 (0.0 -10.0)<br>4.0 ± 2.7 (0.0 -10.0)<br>p-value = 0.06 | 5.6 ± 2.1 (0.0 – 10.0)<br>4.9 ± 2.0 (0.0 – 10.0)<br>p-value = 0.003* |
| Employment (N=248) | No paid work: 101 (40.7%)<br>Paid work: 147 (59.3%) | 4.7 ± 2.9 (0.0 -10.0)<br>3.8 ± 2.7 (0.0 -10.0)<br>p-value = 0.01* | 5.6 ± 2.0 (0.0 – 10.0)<br>4.8 ± 2.0 (0.0 – 10.0)<br>p-value = 0.001* |
| Income (N=291) | < \$41,998: 172 (59.1%)<br>≥ \$ 41,998: 119 (40.9%) | 4.5 ± 2.9 (0.0 -10.0)<br>3.9 ± 2.7 (0.0 -10.0)<br>p-value = 0.08 | 5.5 ± 2.0 (0.0 – 10.0)<br>4.7 ± 2.0 (0.0 – 9.0)<br>p-value = 0.001* |
| Neuro-QoL Stigma (N=243) | 57.4 ± 6.4 (36.0 – 76.9) | $\rho$ = 0.33<br>p-value < 0.0001* | $\rho$ = 0.26<br>p-value < 0.0001* |
| PROMIS Anxiety (N=295) | 57.7 ± 8.9 (32.9 – 84.9) | $\rho$ = 0.31<br>p-value < 0.0001* | $\rho$ = 0.27<br>p-value < 0.0001* |
| PROMIS Depression (N=295) | 55.9 ± 9.5 (34.2 – 79.8) | $\rho$ = 0.33<br>p-value < 0.0001* | $\rho$ = 0.26<br>p-value < 0.0001* |
| PROMIS SRA (N=295) | 45.0 ± 8.0 (22.0 – 68.7) | $\rho$ = -0.45<br>p-value < 0.0001* | $\rho$ = -0.33<br>p-value < 0.0001* |
| Years since pain onset (N=295) | 17.9 ± 13.4<br>(0.1 – 64.3) | $\rho = 0.08$<br>p-value = 0.20 | $\rho = 0.02$<br>p-value = 0.71 |
| Change in best motor function since childhood (N=293) | No: 79 (27.0%)<br>Yes: 214 (73.0%) | 3.3 ± 2.6 (0.0 -10.0)<br>4.6 ± 2.8 (0.0 -10.0)<br>p-value = 0.0002* | 4.6 ± 1.8 (0.0 – 9.0)<br>5.4 ± 2.1 (0.0 – 10.0)<br>p-value = 0.004* |

Note:
\*p<0.05
*GMFCS – Gross Motor Function Classification System*
*Neuro-QOL – Quality of Life in Neurological Disorders*
*PROMIS – Patient-Reported Outcomes Measurement Information System*
*SRA – Satisfaction with Roles and Activities*

The following variables were significantly associated with higher pain intensity: identification as African American race or Hispanic/Latino ethnicity; lower education (up to Associate degree); no current paid employment; lower income; greater levels of stigma, anxiety, and depression; lower levels of satisfaction with social roles and activities; and change in best motor function since childhood since childhood. All other variables did not show significant associations with pain interference or with pain intensity.

Ordinary least squares regression models predicting pain interference and predicting pain intensity are reported in Tables 3a and 3b. Stigma, anxiety, and functional change were found to be significantly positively associated with pain interference (adjusted R^2^=0.17). Hispanic ethnicity, lower income, anxiety, and less satisfaction with social roles and activities were found to be significantly positively associated with pain intensity (adjusted R^2^=0.16).

**Table 3a.** Ordinary least squares regression model with factors associated with pain interference (0-10 point scale; N=238; R2=0.18; Adjusted R2=0.17).

| Explanatory Variable | Parameter Estimate | Standard Error | p-value | Standardized Estimate | 95% Confidence Interval | Variance Inflation Factor |
| --- | --- | --- | --- | --- | --- | --- |
| Neuro-QoL Stigma T-score* | 0.09 | 0.03 | 0.01 | 0.19 | [0.02 – 0.15] | 1.50 |
| PROMIS Anxiety T-score* | 0.06 | 0.02 | 0.01 | 0.20 | [0.02 - 0.11] | 1.51 |
| Change in best motor function since childhood | 1.22 | 0.38 | 0.002 | 0.19 | [0.47 – 1.96] | 1.03 |

\**T-score* = (mean 50, SD 10) based on normative data
*Neuro-QoL – Quality of Life in Neurological Disorders*
*PROMIS – Patient-Reported Outcomes Measurement Information System*

**Table 3b.**
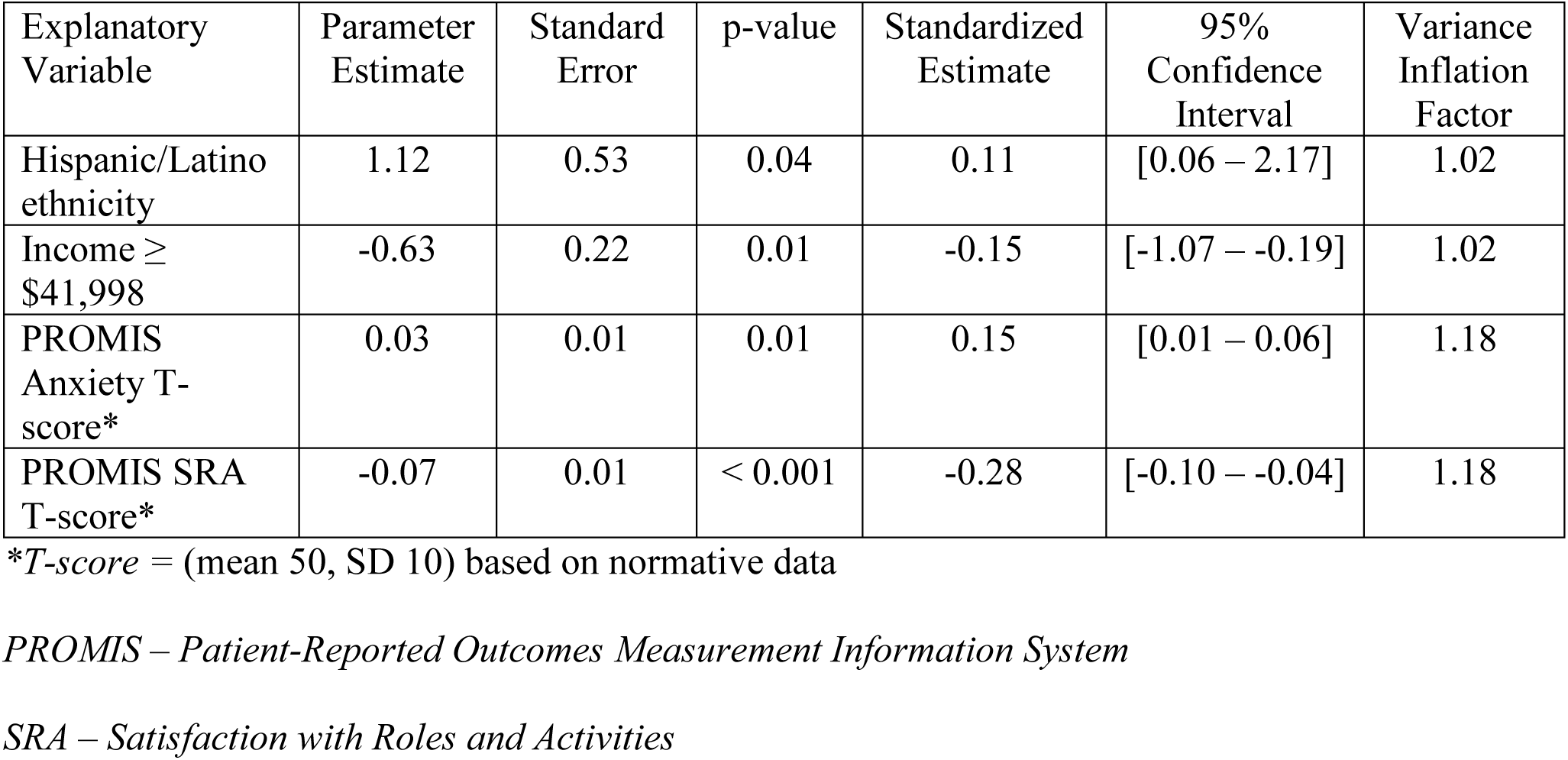
Ordinary Least Squares regression model with factors associated with pain intensity (0-10 point scale; N=290; R2=0.18; Adjusted R2=0.16).

## Discussion

We sought to determine which biopsychosocial factors were associated with pain intensity and pain interference in adults with CP. We found stigma, change in best motor function since childhood, and anxiety best explain pain interference in regression modeling. Additionally, lower income, Hispanic/Latino ethnicity, greater anxiety, and less satisfaction with roles and activities best explained pain intensity. There was no association between duration of pain or ambulation status and pain interference or intensity, contrary to our hypotheses.

This study is the first to evaluate potential relationships between stigma and pain interference in the adult CP population with LBP, which faces the stigmatization of physical disability in addition to the stigmatization of chronic pain. Stigma has been suggested as a barrier to seeking support for managing CP symptoms.(35) Previous studies have shown that stigmatization also adds to the burden of chronic pain in the general chronic pain population.(36) Additionally, stigmatization of disability and chronic pain on the part of healthcare providers can lead to suboptimal patient care, decreased treatment-seeking, and a lack of trust.(37) While we hypothesized that those who had experienced pain for a longer duration might experience less pain interference, and that non-ambulatory status would be associated with lower pain interference, those hypotheses were not supported in our study. Therefore, years since pain onset and ambulatory status would not be considered good indicators of increased pain intensity or interference. This finding is clinically relevant as adults with CP, regardless of ambulatory status and duration of chronic pain, are at risk of disability from LBP. Of interest, adults in our sample identified an average 18 (± 13.4) year history of pain, indicating a need for early screening of chronic LBP.

Physical functional change since childhood was significantly associated with pain interference. Previous literature in children with CP showed a negative effect of pain on gross motor function and participation in recreational activities,(16,38) which could lead to functional decline in adulthood. However, decreasing functional mobility over time could also exacerbate chronic pain and/or its related disability. Additionally, the respondents in this analysis are a subset of larger group of whom more than half self-reported that pain contributed to their functional decline.(22) Physical activity and exercise can mitigate pain intensity and pain interference in people with LBP;(39) and adults with CP with functional decline could benefit from individualized exercise programs that address strength,(40) flexibility,(41) and cardiorespiratory health.(42) Future studies should more closely examine the temporal relationship between pain interference and functional decline, and interprofessional services to optimize function and wellbeing.

Our study found that identifying as Hispanic/Latino increased risk of higher pain intensity. However, one limitation of this study is that our sample demographics overrepresent White women with high levels of education and income, and only 5% of the sample reported Hispanic/Latino ethnicity. Nonetheless, our findings confirm previous literature that has documented a disparity in pain intensity and interference, with higher levels among people who are non-White and have low levels of education and income, across different types of populations with pain.(13) This emphasizes the critical role that psychological and socioenvironmental factors play, and highlights the importance of assessing and addressing these factors in clinical practice.

Anxiety emerged as significant in the models for both pain interference and pain intensity. Adults with CP have higher rates of psychiatric disorders and are more likely to experience anxiety than age, sex, and practice-matched controls.(43) Anxiety disorders and pain interference have been shown to be closely related,(44) and the presence of pain interference can decrease responsiveness to treatment for anxiety disorders.(45) Therefore, screening and treatment for anxiety are critical to optimizing care for adults with CP and chronic pain.

### Limitations

This study sample represents self-selected survey respondents with internet access and generally higher education, which could decrease the generalizability of our findings. Also, all of our measures were self-reported without clinical data, which can lead to a risk of bias. Therefore, the generalizability of our results is limited to individuals with similar characteristics. The cross-sectional nature of this study is a further limitation as we could not assess longitudinal relationships. The study is also limited by examining only person related factors. Issues such as rural versus urban or specialty versus primary care, number of specialists seen, transportation, knowledge of providers, and family support systems were not examined. Also, separate analysis of the outcomes for those requiring more time or assistance for communication was not performed. Future studies should identify modifiable factors that may reduce disparities in service use and outcomes.

## Conclusions

This study suggests that psychosocial health, ethnicity, stigma, and functional change all interplay with chronic low back pain in adults with CP. Psychological and social factors, as well as physical function decline, were significantly associated with pain interference and pain intensity. Therefore, these factors should be considered in a comprehensive evaluation and management plan for LBP in adults with CP. Future research should 1) identify potential targets for interventions to reduce functional decline throughout the CP lifespan in the context of chronic pain, 2) further evaluate mechanisms of stigma and its impact on chronic pain, and 3) further evaluate social determinants of health such as race, ethnicity, education, and income to stratify risk for development of chronic pain and associated disability in adults with CP.

## Data Availability

Research data are not shared.

